# Real-world study of intranasal ketamine for use in patients with refractory chronic migraine

**DOI:** 10.1101/2022.11.20.22282558

**Authors:** Hsiangkuo Yuan, Aniket Natekar, Jade S. Park, Clinton Lauritsen, Eugene R. Viscusi, Michael J. Marmura

## Abstract

Subanesthetic ketamine infusion has been used for managing refractory headache in inpatient or outpatient infusion settings. Intranasal (IN) ketamine may be an alternative option for outpatient care.

We performed a retrospective study at a single tertiary headache center to assess the clinical effectiveness and tolerability of IN ketamine in patients with refractory chronic migraine (rCM). Candidates who received IN ketamine between January 2019 and February 2020 were screened through an electronic medical record query. Manual chart reviews and structured phone interviews were conducted upon obtaining informed consent.

Among 242 subjects screened, 169 (age 44.3±13.8; female 79.9%) were interviewed. They reported 25.0±8.7 monthly headache days and tried 6.9±3.1 preventive medications. Overall, they used roughly 7.8±7.0 sprays (ie., 78 mg) per day and 11.6±8.9 days per month. Intranasal ketamine was reported as “very effective” in 49.1% and quality of life (QOL) was considered “much better” in 35.5%. However, 74.0% reported at least one adverse event (AE).

In this retrospective study, IN ketamine can serve as an acute treatment for rCM by reducing headache intensity and improving QOL with relatively tolerable AEs. Most patients found IN ketamine effective and continued to use it despite these AEs. The study is limited by its single-center design and selection/recall biases. Well-designed prospective placebo-controlled trials are necessary to demonstrate the efficacy and safety of IN ketamine in patients with migraine.

- **What is already known on this topic –** Intravenous ketamine, although has been used for chronic pain and refractory headache, is limited to infusion settings. Intranasal ketamine, a more convenient alternative, has not been well-studied for refractory headache.
- **What this study adds –** This real-world study describes the usage pattern, effectiveness, and adverse event profiles of intranasal ketamine in patients with refractory chronic migraine.
- **How this study might affect research, practice or policy –** Intranasal ketamine is probably effective with minimal adverse events for refractory chronic migraine, but more well-designed studies are needed.

## INTRODUCTION

Refractory chronic migraine (rCM), defined by an inadequate response to multiple proven preventive and acute medications with a significant impact on disability and quality of life (QOL),^1^ is highly pervasive in tertiary headache clinics. Ketamine, a dissociative anesthetic agent, has potential utility for perioperative pain, chronic pain, depression, and headache,^2,3^ especially when used with benzodiazepines to mitigate psychomimetic AEs.^4^ Known as a non-competitive N-Methyl-D-aspartate (NMDA) receptor antagonist, ketamine inhibits nitric oxide synthase, proinflammatory cytokine release, and serotonin reuptake. It antagonizes voltage-gated sodium channels, large-conductance potassium channels, L-type voltage-dependent calcium channels, calcitonin gene-related peptide (CGRP) receptors, and nicotinic acetylcholine receptors. Ketamine also activates μ/δ opioid receptors, AMPA receptors, and GABA_A_ receptors, and upregulates brain-derived neurotrophic factor.^3,5-7^ There are two stereoisomers: S(+) and R(-), where S(+) isomer is 3- to 4-fold more potent than R(-) but has quicker clearance. The mechanism of action for ketamine in antinociception is likely multifactorial beyond NMDA but remains unclear.^8^

To date, several randomized control trials (RCTs) have been published on subanesthetic ketamine infusion for headache management.^9^ Since 2000, Thomas Jefferson University Hospital has been using subanesthetic ketamine infusions for multiple chronic pain conditions, including chronic regional pain syndrome and headache. We published several retrospective cohort studies demonstrating ketamine infusion’s potential utility in managing rCM.^10-13^

However, intravenous (IV) ketamine typically requires dose titration and AE monitoring by a pain specialist in the hospital, thus limiting its use in the outpatient setting. IN ketamine, with its simple storage and convenient administration, is an attractive alternative to IV administration. It has rapid systemic absorption (time of onset 5-10 minutes) without first-pass hepatic metabolism but with 25-50% bioavailability (higher than oral ketamine). The FDA has recently approved esketamine (Spravato; Janssen, Raritan, NJ), an IN S (+) enantiomer for treatment-resistant depression; its use in treating headache or migraine has not been studied.

Over the last few years, IN ketamine has been evaluated for acute pain management.^14,15^ While there is some evidence behind IN ketamine as a treatment for headache disorders (e.g., migraine, cluster headache, non-traumatic headache) in several studies,^16-21^ its role in treating rCM has not been well described nor validated. At our center, IN ketamine is often prescribed to patients with rCM who do not respond well to standard infusion treatments, including dihydroergotamine and lidocaine, before or after a scheduled ketamine infusion. We hypothesized that IN ketamine alleviates acute headaches in patients with rCM as adjunctive therapy to their standard-of-care headache management. To better understand the real-world benefit of IN ketamine, we retrospectively reviewed the effectiveness and tolerability of IN ketamine in patients with rCM as outpatients at a tertiary headache center.

## METHODS

This single-center study was approved by the Thomas Jefferson University Institutional Review Board (#20E.147). It involves retrospective chart reviews and phone interviews with established patients at the Jefferson Headache Center (JHC). Patients who did not have success with multiple standard migraine treatments were offered IN ketamine scripts (100mg/ml, 30ml), which were formulated by a local compounding pharmacy to approximately 10mg per 0.1ml spray and instructed to use 1-3 sprays each nostril per dose at the discretion of their JHC providers. IN ketamine safety precautions were reviewed with patients to decrease the risk of addiction and misuse. All patients given IN ketamine scripts were required to sign a treatment contract agreeing to frequent follow-up visits, avoid use before driving, refrain from using other controlled substances without our knowledge or alcohol, and avoid pregnancy.

We identified patients who received IN ketamine scripts between 1/1/2019 to 2/29/2020 through an electronic medical record query. Eligible patients were at least 18 years of age at screening and received at least one electronic script for IN ketamine during the study period. All identified subjects were mailed a recruitment letter regarding the study, its goals, and the option to opt-out. After a 60-day waiting period, identified patients were contacted by phone to conduct a structured interview upon obtaining verbal consent. Participants were excluded if they never filled the ketamine script, received IN ketamine for non-migraine diagnoses, or could not participate via phone interview.

Two independent investigators performed chart reviews involving demographic information, headache diagnoses, comorbidities, headache characteristics, current and past preventive regimens, medication overuse at the time of review, and the setting and reason for IN ketamine initiation. Through phone interviews, we collected data including current IN ketamine regimen (dose, frequency), time to pain relief, consistency of pain relief, changes in pain level (11-point numerical rating scale) before and after use, global impression of effectiveness, the overall impact on QOL, pain catastrophizing scale, and self-efficacy scale and AEs. The collected information was stored on a HIPAA-compliant web-based REDCap electronic data capture tool hosted at Thomas Jefferson University.^22^ Upon data cleaning and verification, the senior investigator determined final data approval.

### Statistical Analysis

De-identified data were analyzed using the statistical analysis program SPSS Statistics (v.28, IBM, Armonk, NY). Descriptive data were presented as percentages or arithmetic means ± standard deviations and medians (interquartile ranges). Regression analyses were performed using the Generalized Linear Models (GLM) to describe the association between dependent variables (pain intensity change [continuous variables], overall impression as very effective vs. others [binary variables]) and multiple key headache characteristics (daily headache, pain catastrophizing score, self-efficacy score, previous positive response to IV ketamine). Variables were fit independently as main effects controlling for age, sex, and body-mass index (BMI). Odds ratios and β coefficients were reported for binary or continuous variables, respectively, where p-values <0.05 were considered significant. Missing data were considered at random, and no imputation was performed.

## RESULTS

In total, 242 rCM patients were prescribed IN ketamine, and 169 were successfully contacted, consented, and interviewed. The remaining 55 patients could not be reached, and 18 declined participation. Table 1 illustrates the demographics of the study population. While all patients had a migraine diagnosis (with 38.5% having aura), coexisting headache diagnoses included new-daily persistent headache (n=22, 13.0%), post-traumatic headache (n=8, 17.8%), and idiopathic intracranial hypertension (n=5, 3.0%). Common comorbidities included psychiatric disorders (e.g., anxiety, depression, bipolar, PTSD) and other somatic symptoms (e.g., back, neck, TMJ pain). The participants were mostly Caucasian (161, 95.3%), female (79.9%), 44.3±13.8 years old, with a BMI of 30.36±7.78, and college/graduate education (56.2%). They reported 25.0±8.7 monthly headache days (MHD) and 13.7±10.2 monthly disabling headache days, where more than two-thirds (67.5%) had daily headaches. Forty-four patients (45.6%) were on employment disability. On average, they tried 6.9±3.1 preventive medications in the past and were on 2.6±1.7 preventive medications at the time of the interview. 143 (84.6%) and 38 (22.5%) tried more than 3 or 5 classes of preventive medications. While 101 (59.8%) and 86 (50.9%) previously tried CGRP mAbs and onabolulinumtoxinA, 89 (52.7%) and 73 (43.2%) were on these treatments, respectively, at the time of the interview. For acute treatment, the most prescribed medications were ketorolac injection (51, 30.2%), and dihydroergotamine injection (41, 24.3%). The pain catastrophizing scale and self-efficacy scale were 4.73±3.33 and 7.96±2.28, respectively, at the time of the interview.

**Table 1.** Descriptive of the study population

| Table 1. Descriptive of the study population |  |
| --- | --- |
| Demographics |  |
| Caucasian, n (%) | 161 (95.3) |
| Female, n (%) | 135 (79.9) |
| Age (year) | 44.33±13.77 (18-75), 43 (22) |
| BMI | 30.36±7.78 (15.1-57.8), 29.6 (8.75) |
| Aura, n (%) | 65 (38.5) |
| Headache characteristics |  |
| Migraine years | 12.05±9.92 (0-50), 8.5 (13) |
| Current monthly headache days | 25.00 ± 8.68 (0-30), 30 (9) |
| Daily headache, n (%) | 111 (65.7) |
| Current monthly bad HA days | 15.13±9.93 (0-30), 15 (19) |
| Current monthly disabled days | 13.67±10.23 (0-30), 13 (17) |
| On employment disability, n (%) | 77 (45.6) |
| No. of current preventives | 2.62±1.70 (0-7), 3 (3) |
| No. of previous preventives | 6.89±3.07 (0-16), 7 (4) |
| Pain Catastrophizing Scale | 4.73±3.33 (0-12), 4 (5) |
| Self-Efficacy Scale | 7.96±2.28 (0-10), 9 (3) |
| Comorbidity |  |
| Neck pain, n (%) | 103 (60.9) |
| Anxiety, n (%) | 99 (58.6) |
| Depression, n (%) | 93 (55.0) |
| Insomnia, n (%) | 74 (43.8) |
| Low back pain, n (%) | 68 (40.2) |
| Mild concussion, n (%) | 58 (34.3) |
| TMJ disorder, n (%) | 39 (23.1) |
| Hypothyroidism, n (%) | 26 (15.4) |
| PTSD, n (%) | 25 (14.8) |
| OSA, n (%) | 23 (13.6) |
| Bipolar, n (%) | 16 (9.5) |
| Data are presented in case number (percentage) or mean ± standard deviation (range), median (interquartile range).<br>Abbreviations: BMI: body mass index. HA: headache. TMJ: temporomandibular joint. PTSD: post-traumatic stress disorder. OSA: obstructive sleep apnea | |

One hundred ten patients (65.1%) were current IN ketamine users. Forty-one (24.7%) and forty-six (27.7%) patients were offered IN ketamine before and after ketamine infusion, respectively. 47.6% had never received ketamine infusion. The most common reasons for initiating IN ketamine were an incomplete response to prior acute medications (100, 59.2%), incomplete response to prior preventives (52, 30.8%), prior benefit from IV ketamine (38, 22.5%), and unsuccessful lidocaine infusion (22, 13.0%). Concomitant acute medications included NSAIDs (n=85, 50.3%), DHE (n=52, 30.8%), triptans (n=34, 20.1%), neuroleptics (n=60, 35.5%), opioids (n=21, 12.4%), gepants (n=35, 20.7%), simple analgesics (n=22, 13.0%), and cannabinoids (n=20, 11.8%). Table 2 shows the effectiveness metrics of IN ketamine. Overall, patients used roughly 7.8 sprays per day and 12 days per month. Twenty-three (13.6%) were daily users, and 31 (18.3%) used more than 10 sprays per day. Nine used it as preventive, and 11 for the aura. There was a decrease of 2.87±1.95 (within group p<0.001) in pain intensity after using IN ketamine. The average time for onset of pain relief was 73.63 minutes, with 70.6% treatment response consistency. Compared to other acute medications, IN ketamine was reported as much better in 43.2% (n=73), somewhat better in 29.6% (n=50), and no different or worse in 22.6% (n=38). One hundred twenty patients (71.0%) reported using fewer acute medications. When evaluating overall effectiveness, 83 (49.1%) found it very effective, 67 (39.6%) somewhat effective, 16 (9.5%) reported no change, and 3 (1.8%) reported worse than prior acute medications. Within the same group, 60 (35.5%) found the overall impact of IN ketamine on their QOL to be much better, 72 (42.6%) found it to be somewhat better, 31 (18.3%) reported no change, and 6 (3.6%) worse.

**Table 2.**
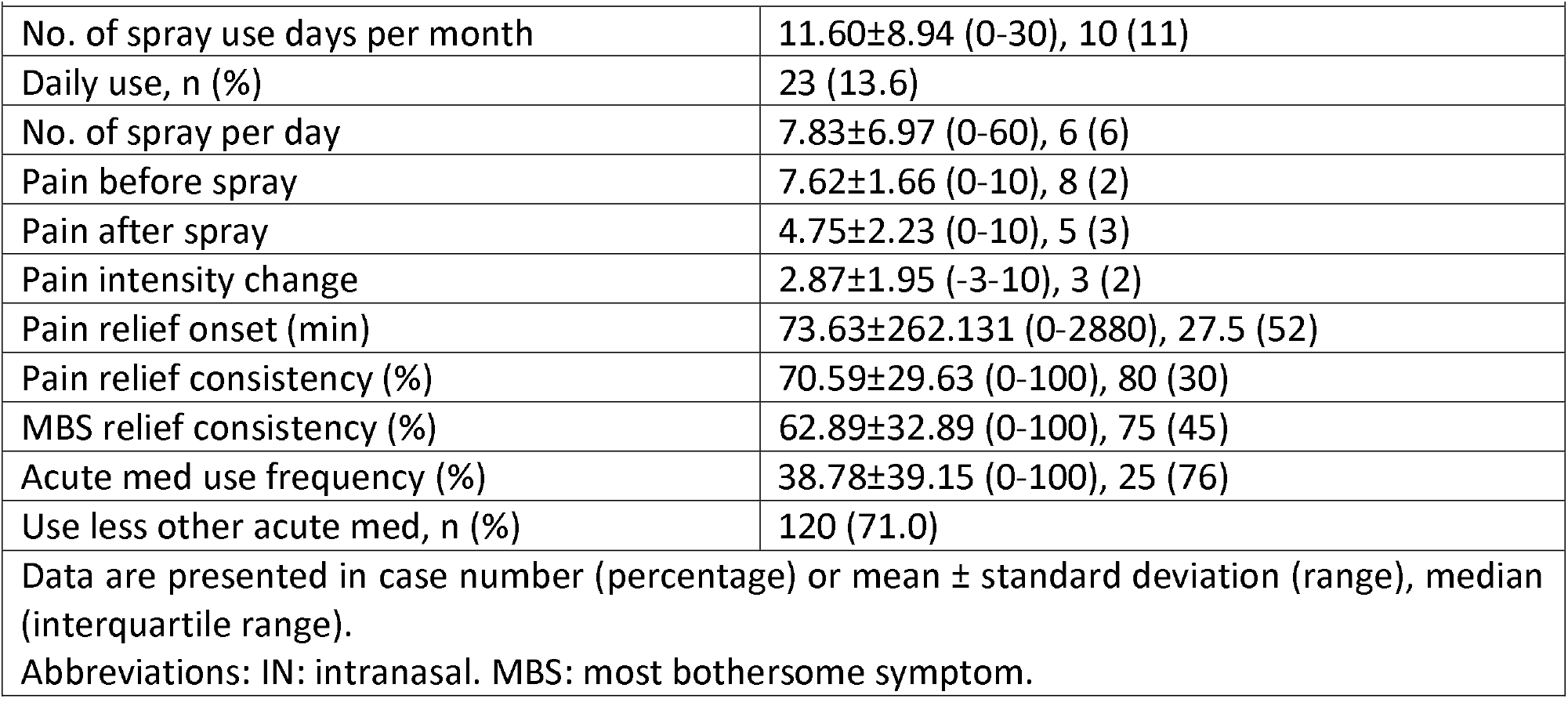
Evaluation of IN ketamine effectiveness

| **Table 2.** Evaluation of IN ketamine effectiveness |  |
| --- | --- |
| No. of spray use days per month | 11.60±8.94 (0-30), 10 (11) |
| Daily use, n (%) | 23 (13.6) |
| No. of spray per day | 7.83±6.97 (0-60), 6 (6) |
| Pain before spray | 7.62±1.66 (0-10), 8 (2) |
| Pain after spray | 4.75±2.23 (0-10), 5 (3) |
| Pain intensity change | 2.87±1.95 (-3-10), 3 (2) |
| Pain relief onset (min) | 73.63±262.131 (0-2880), 27.5 (52) |
| Pain relief consistency (%) | 70.59±29.63 (0-100), 80 (30) |
| MBS relief consistency (%) | 62.89±32.89 (0-100), 75 (45) |
| Acute med use frequency (%) | 38.78±39.15 (0-100), 25 (76) |
| Use less other acute med, n (%) | 120 (71.0) |
| Data are presented in case number (percentage) or mean ± standard deviation (range), median (interquartile range).<br>Abbreviations: IN: intranasal. MBS: most bothersome symptom. | |

We evaluated pain intensity change and overall impression (very effective vs. others) using GLM adjusted for age, sex, and BMI. Demographics (e.g., age, sex, BMI) were not significantly associated with pain intensity change or very effective response. The very effective overall impression and pain intensity change were found to be significantly associated with the current use of IN ketamine (odds ratio of 5.601 [2.436-10.5] and 1.908 [1.029-3.539]) but not others (Table 3).

**Table 3.** Regression analyses for IN ketamine effectiveness

| **Table 3.** Regression analyses for IN ketamine effectiveness |  |  |  |
| --- | --- | --- | --- |
|  | Pain intensity change | Very effective vs. others | P value |
| Current IN ketamine user | 1.908 (1.029-3.539) | 5.061 (2.436-10.5) | P=0.040, P<0.001 |
| Daily HA | 0.613 (0.328-1.144) | 1.557 (0.796-3.044) | P=0.124, P=0.196 |
| # ketamine use day/month | -0.010 (-0.043-0.023) | 0.032 (-0.004-0.068) | p=0.535, p=0.079 |
| # ketamine spray per day | 0.039 (-0.003-0.081) | 0.014 (-0.031-0.059) | P=0.068, p=0.556 |
| Positive IV ketamine response | 1.155 (0.570-2.342) | 1.881 (0.880-4.018) | p=0.404, p=0.103 |
| Pain catastrophizing score | -0.013 (-0.101-0.075) | -0.053 (-0.146-0.040) | p=0.769, p=0.265 |
| Self-efficacy score | 0.018 (-0.112-0.147) | 0.018 (-0.118-0.153) | p=0.790, p=0.800 |

Independent variables were analyzed individually by the generalized linear models (linear or binary logistic) adjusted for age, sex, and BMI. Data were presented as $\beta$ coefficient (95% CI) for continuous variables and odds ratio (95% CI) for binary variables.
Abbreviation: CI: confidence interval. IN: intranasal. IV: intravenous.

Table 4 lists the AEs reported by the participants. One hundred twenty-five patients (74.0%) reported at least one AE. Fatigue and double vision/blurred vision were the most common, followed by cognitive AEs (e.g., confusion/dissociation, vivid dreams, hallucination). Reported nausea and dizziness were expected, as known AEs from ketamine. In addition, among 142 participants with laboratory data, alanine and aspartate transaminase elevations (>3x upper normal limit) were found in 4.9% (n=7) and 2.1% (n=3), respectively, without bilirubin elevation. Short-term transaminase elevations were discovered during inpatient ketamine infusion (n=4), while others were due to medical issues (e.g., fatty liver, gallstones). KNS usage continued afterward without complication.

**Table 4.** Reported adverse events by surveyed patients

| **Table 4.** Reported adverse events by surveyed patients |  |
| --- | --- |
| Presence of $\geq 1$ adverse events, n (%) | 125 (74.0) |
| Fatigue, n (%) | 37 (21.9) |
| Double/blurred vision, n (%) | 36 (21.3) |
| Confusion/dissociation, n (%) | 34 (20.7) |
| Nausea, n (%) | 28 (16.6) |
| Dizziness, n (%) | 23 (13.6) |
| Nasal discomfort/epistaxis, n (%) | 21 (12.4) |
| Vivid dreams, n (%) | 17 (10.1) |
| Hallucination, n (%) | 13 (7.7) |
| Ageusia, n (%) | 10 (5.9) |
| Increased anxiety, n (%) | 6 (3.6) |
| Vomiting, n (%) | 5 (3.0) |
| Tremor, n (%) | 5 (3.0) |
| Imbalance, n (%) | 5 (3.0) |
| Worsened/rebound headache, n (%) | 4 (2.4) |
| Data presented in case number and percentage of total patients |  |

## DISCUSSION

In this retrospective single-center study with 169 interviewed participants, mostly with rCM who were unsuccessful with multiple preventive medications and biologics, adjunctive IN ketamine significantly improved acute headache pain intensity with consistent treatment response and reductions in other acute medication use. This is the first outpatient IN ketamine study for rCM. Almost half of the participants considered IN ketamine very effective, and more than two-thirds of patients found it improved their QOL, especially among current active users. Variables such as daily headache, number of IN ketamine spray/day and day/month, previous response to ketamine infusion, pain catastrophizing score, and self-efficacy score were not associated with pain intensity change or overall effectiveness. Although almost three-quarters reported having at least one AE, particularly fatigue, vision disturbances, and cognitive issues, many continued using it.

To date, there are only a limited number of studies using IN ketamine for headache treatment (Table 5). In 2000, Kaube et al. presented a case series of 11 patients with familial hemiplegic migraine who self-administered 25mg of IN ketamine. Five showed reduced severity and duration of the neurological deficit.^16^ In 2013, Afridi et al. reported the first double-blind, randomized controlled trial (DBRCT) comparing IN ketamine 25mg against IN midazolam 2mg in 18 patients with migraine with prolonged aura. IN ketamine reduced the severity (p=0.032) but not the duration of the aura, whereas midazolam had no effect.^17^ In the THINK trial (single-blind RCT), Benish et al. compared IN ketamine (0.75 mg/kg) vs. IN ketamine (0.015 mg/kg) + IV metoclopramide (10mg) + oral diphenhydramine (25mg) in 53 patients with primary headache syndrome in the emergency department (ED). The average change in pain visual analog scale (VAS) at 30 min post-intervention (the primary endpoint) was 22.2 mm in the control arm vs. 29.0 in the IN ketamine arm (effect size difference 6.8 mm, 95% CI -5.8 to 19.4); no statistically significant difference.^18^ Recently, Sarvari et al., in a DBRCT investigated the efficacy of IN ketamine (0.75 mg/kg) vs. IV ketorolac (30mg) in 140 patients with non-traumatic acute headache in the ED. Pain reduction was significantly greater in IN ketamine than in IV ketorolac at 30, 60, and 120 min.^21^ These RCTs showed that IN ketamine may be as effective as the standard IV headache regimen. Apart from RCTs, Turner et al. retrospectively reviewed 34 patients with status migrainosus receiving IN ketamine (0.1-0.2 mg/kg) in an inpatient setting. Twenty-five (73.5%) were responders with an average pain score reduction of -7.2 from admission to discharge,^19^ suggesting potential utility in severe migraine. Petersen et al. also showed that IN ketamine 15mg every 6 minutes (max 5 uses) could reduce pain intensity of 1.1 (95%CI −0.6 to 2.7) in 15 minutes and 4.3 (95%CI 2.4-6.2) in 30 minutes among 20 cluster headache patients.^20^ Esketamine, a S-enantiomer of ketamine, was FDA-approved for treatment-resistant depression. Considering that depression and anxiety are significant comorbidities associated with headache disorders, IN ketamine could potentially treat depression in patients with rCM. It is important to know that all studies reported AEs, including dizziness, nausea, increased BP/HR, fatigue, and mood change. While these AE were expected, they were temporary and resolved within a few hours. Based on these studies, IN ketamine appeared to reduce head pain quickly and effectively, at least for common headaches in the ED. Our study, which included mostly patients with rCM, further expands the potential utility of IN ketamine in the refractory headache population.

**Table 5.** Summary of IN ketamine trials for headache treatment

| **Table 5.** Summary of IN ketamine trials for headache treatment |  |  |  |
| --- | --- | --- | --- |
| Study | Design | Ketamine dose | Primary findings |
| 2000, Kaube et al.<sup>16</sup> | Case series. Familial hemiplegic migraine. N=11 | 25mg, self-medication | IN ketamine reproducibly reduced the severity and duration of the neurologic deficits in 5 patients |
| 2013, Afridi et al.<sup>17</sup> | DBRCT. Migraine with prolonged aura. N=18 | 25mg vs. 2mg IN midazolam | IN ketamine reduced the severity ($p=0.032$) but not duration of aura, whereas midazolam had no effect. |
| 2019, Benish et al.<sup>18</sup><br>NCT03081416. | SBRCT. Primary headache syndrome. N=53 | 0.75 mg/kg ± 0.25 mg/kg vs. 0.015 mg/kg ketamine + 10mg IV metoclopramide + 25mg diphenhydramine | The mean change in pain VAS score at 30 min post intervention was 22.2 mm in the control arm vs. 29.0 mm in the IN ketamine arm (effect size difference 6.8 mm, 95% CI -5.8–19.4) |
| 2020, Turner et al.<sup>19</sup> | Retrospective. Status migrainosus. N=34 | 0.1-0.2mg/kg, inpatient | 25 (73.5%) were responders with mean pain score reduction of -7.2 from admission to discharge. 66.1% pain reduction overall. |
| 2022, Petersen et al.<sup>20</sup><br>NCT04179266 | Open label. CH.<br>N=20 | 15mg every 6 minutes<br>(max 5 times) | After 15 min, pain intensity reduction was 1.1 (95%CI -0.6 to 2.7). After 30 min, reduction was 4.3 (95%CI 2.4-6.2). |
| 2022, Sarvari et al.<sup>21</sup><br>IRCT20180108038276N3 | DBRCT. Non-traumatic acute headache. N=140 | 0.75mg/kg vs. 30mg IV ketorolac | Pain reduction was significantly greater in IN ketamine than IV ketorolac at 30, 60, 120 min. |
| Abbreviation: DBRCT: double-blind randomized controlled trial. SBRCT: single-blind randomized controlled trial. VAS: visual analog scale. CH: cluster headache. |  |  |  |

Since ketamine has a relatively short half-life (about 2 hrs), refractory headache patients may tend to use it more regularly despite receiving counseling. In our population, 23 (13.6%) used it daily, 37 (21.9%) at least 15 days/month, and 76 (45.0%) at least 10 days/month. Although, the usage frequency was not associated with pain intensity change or overall effectiveness. While daily headache is generally considered more refractory to treatment, there was no difference in the usage frequency between those with or without daily headache (12.03±8.44 vs. 10.75±9.90 days/month, p=0.387; 7.4±5.19 vs. 8.31±9.64 sprays/day, p=0.428). Even though almost three-quarters of the participants reported AEs, many continued to use IN ketamine and reported better responses than other abortive medications. The active metabolite of ketamine, hydroxynorketamine, has a much longer half-life than that of ketamine and lacks the addictive effect.^23,24^ Hydroxynorketamine provides an additional mechanism of action; it blocks NMDA receptor currents with low affinity and weak voltage dependence, and it is effective when applied to resting receptors.^25^ It also elicits antidepressant effects via the inhibition of AMPA glutamate and α7 nicotinic cholinergic receptors.^5^ With hydroxynorketamine’s longer half-life, the frequent use of ketamine may lead to the accumulation of hydroxynorketamine to provide better pain inhibition, as seen in the ketamine infusion study.^12^ However, the optimal dosage for IN ketamine that is safe and effective remains to be determined.

To date, guidelines around the optimal dose of IN ketamine are lacking. IN ketamine has a Tmax of 10-20 minutes and a wide bioavailability of 8-45%.^26^ However, such metrics were gathered from blood and may not reflect ketamine’s actual distribution in the trigeminal system. It is important to understand that IN absorption varies by the nasal spray apparatus, nasal passage, site of deposition, spray viscosity, and other factors. In our study, ketamine was delivered via a traditional metered-dose spray pump with 100 μL (10mg) per spray (without any mucoadhesive or permeabilization agent), generating particles between 50-100 μm in diameter. Some studies used MAD Nasal™, which is an atomization device that produces smaller particles (30-100 μm) and delivers deeper/higher into the nasal cavity and less to the lung than the typical nasal spray.^27,28^ Compared to the lower nasal space, its upper counterpart allows for more efficient absorption due to looser olfactory epithelium tight function, lower mucociliary clearance, and richer vascular/lymphatic system.^29^ Lipophilic small molecules such as ketamine can be transported via transcellular, paracellular, and perineural pathways via trigeminal nerves,^29,30^ offering a possible delivery route to the trigeminal ganglia bypassing the blood-ganglion barrier. It is plausible that the local concentration within the trigeminal system may be higher than the serum concentration, creating a localized antinociceptive effect with lower systemic AE. Even though our participants used an average of 78mg per day, which is much lower than the infusion daily dose (0.5-1 mg/kg/hr; 960-1920 mg daily for an 80 kg person), the cephalic analgesic effect and psychometric AEs were still apparent. At this time, without a specific biomarker reflecting ketamine’s local concentration, the optimal dosage may require individual titration to find a good balance between safety and efficacy. At Jefferson, we routinely ask patients to titrate from a lower dose and limit the IN ketamine use for their headache (not other somatic pain) to a maximum of 20 sprays a day and 40 sprays a week. Participants were instructed to cautiously try different dosages while monitoring ketamine’s potential AEs to avoid overuse or intoxication.

### Strengths and weaknesses

Our study is the first to evaluate IN ketamine as an outpatient acute treatment for rCM in adults. The data was collected through chart review and structured interviews, offering cleaner and more detailed information than chart review alone. However, this study was based on a single tertiary headache center, with the study population limited to mostly Caucasian women. Consequently, the study results may have limited generalizability. In manual chart reviews and phone interviews, some information may not be available or may be limited due to selection and recall biases. Since this study is not a double-blind placebo-controlled trial, the level of evidence is limited regarding clinical management recommendations. Most participants used IN ketamine concomitantly with other abortive and preventive medications. Therefore, it is challenging to assess the therapeutic benefit of IN ketamine in isolation. Regarding IN ketamine’s safety profile, many AEs overlapped with migraine-associated symptoms, including but not limited to nausea and blurry vision. The true safety profile should be further evaluated in a placebo-controlled trial.

This retrospective study suggests that IN ketamine can be a quick, effective, and relatively well-tolerated analgesic for rCM in the outpatient setting. IN delivery may allow for more rapid access of ketamine to the trigeminal system beyond circulation. The cumulative effect of ketamine’s active metabolite, hydroxynorketamine, may further enhance its analgesic impact. The optimal IN ketamine dosage, however, remains to be explored. While IN ketamine has been approved for treatment-resistant depression, well-designed prospective placebo-controlled trials involving IN ketamine for acute treatment of migraine are still needed.

**Figure 1.**
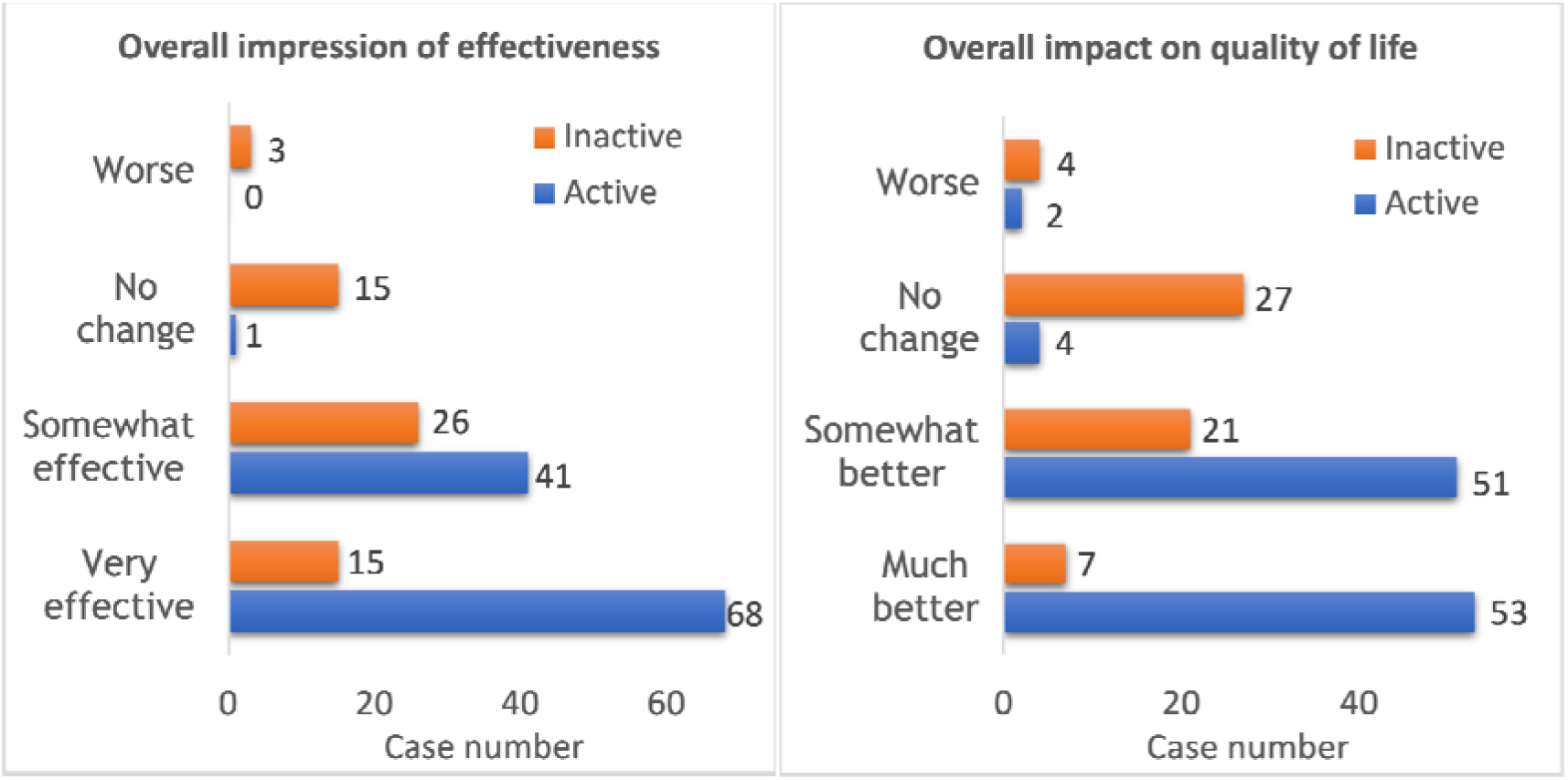
Comparison between inactive and active IN ketamine users regarding overall impression of effectiveness and impact on QOL. Among active users, the majority found IN ketamine as very effective with much better overall impact on QOL.

## Data Availability

All data produced in the present study are available upon reasonable request to the authors

## REFERENCES

1. Sacco S, Braschinsky M, Ducros A, et al. European headache federation consensus on the definition of resistant and refractory migraine : Developed with the endorsement of the European Migraine & Headache Alliance (EMHA). J Headache Pain. 2020;21(1):76. doi:10.1186/s10194-020-01130-5

2. Visser E, Schug SA. The role of ketamine in pain management. Biomed Pharmacother. 2006;60(7):341–348. doi:10.1016/j.biopha.2006.06.021

3. Cohen SP, Bhatia A, Buvanendran A, et al. Consensus Guidelines on the Use of Intravenous Ketamine Infusions for Chronic Pain From the American Society of Regional Anesthesia and Pain Medicine, the American Academy of Pain Medicine, and the American Society of Anesthesiologists. Reg Anesth Pain Med. 2018;43(5):521–546. doi:10.1097/AAP.0000000000000808

4. Green SM, Krauss B. The taming of ketamine-40 years later. Ann Emerg Med. 2011;57(2):115–116. doi:10.1016/j.annemergmed.2010.09.021

5. Zanos P, Moaddel R, Morris PJ, et al. Ketamine and Ketamine Metabolite Pharmacology: Insights into Therapeutic Mechanisms. Pharmacol Rev. 2018;70(3):621–660. doi:10.1124/pr.117.015198

6. Chan KY, Gupta S, de Vries R, et al. Effects of ionotropic glutamate receptor antagonists on rat dural artery diameter in an intravital microscopy model. Br J Pharmacol. 2010;160(6):1316–1325. doi:10.1111/j.1476-5381.2010.00733.x

7. Choudhury D, Autry AE, Tolias KF, Krishnan V. Ketamine: Neuroprotective or Neurotoxic? Front Neurosci. 2021;15:672526. doi:10.3389/fnins.2021.672526

8. Schwenk ES, Pradhan B, Nalamasu R, et al. Ketamine in the Past, Present, and Future: Mechanisms, Metabolites, and Toxicity. Curr Pain Headache Rep. 2021;25(9):57. doi:10.1007/s11916-021-00977-w

9. Chah N, Jones M, Milord S, Al-Eryani K, Enciso R. Efficacy of ketamine in the treatment of migraines and other unspecified primary headache disorders compared to placebo and other interventions: a systematic review. J Dent Anesth Pain Med. 2021;21(5):413–429. doi:10.17245/jdapm.2021.21.5.413

10. Pomeroy JL, Marmura MJ, Nahas SJ, Viscusi ER. Ketamine Infusions for Treatment Refractory Headache. Headache. 2017;57(2):276–282. doi:10.1111/head.13013

11. Schwenk ES, Dayan AC, Rangavajjula A, et al. Ketamine for Refractory Headache: A Retrospective Analysis. Reg Anesth Pain Med. 2018;43(8):875–879. doi:10.1097/AAP.0000000000000827

12. Schwenk ES, Torjman MC, Moaddel R, et al. Ketamine for Refractory Chronic Migraine: An Observational Pilot Study and Metabolite Analysis. J Clin Pharmacol. 2021;61(11):1421–1429. doi:10.1002/jcph.1920

13. Shafqat R, Flores-Montanez Y, Delbono V, Nahas SJ. Updated Evaluation of IV Dihydroergotamine (DHE) for Refractory Migraine: Patient Selection and Special Considerations. J Pain Res. 2020;13:859–864. doi:10.2147/JPR.S203650

14. Li X, Hua GC, Peng F. Efficacy of intranasal ketamine for acute pain management in adults: a systematic review and meta-analysis. Eur Rev Med Pharmacol Sci. 2021;25(8):3286–3295. doi:10.26355/eurrev_202104_25738

15. Seak YS, Nor J, Tuan Kamauzaman TH, Arithra A, Islam MA. Efficacy and Safety of Intranasal Ketamine for Acute Pain Management in the Emergency Setting: A Systematic Review and Meta-Analysis. J Clin Med. 2021;10(17):3978. doi:10.3390/jcm10173978

16. Kaube H, Herzog J, Kaufer T, Dichgans M, Diener HC. Aura in some patients with familial hemiplegic migraine can be stopped by intranasal ketamine. Neurology. 2000;55(1):139–141. doi:10.1212/wnl.55.1.139

17. Afridi SK, Giffin NJ, Kaube H, Goadsby PJ. A randomized controlled trial of intranasal ketamine in migraine with prolonged aura. Neurology. 2013;80(7):642–647. doi:10.1212/WNL.0b013e3182824e66

18. Benish T, Villalobos D, Love S, et al. The THINK (Treatment of Headache with Intranasal Ketamine) Trial: A Randomized Controlled Trial Comparing Intranasal Ketamine with Intravenous Metoclopramide. J Emerg Med. 2019;56(3):248–257 e241. doi:10.1016/j.jemermed.2018.12.007

19. Turner AL, Shandley S, Miller E, Perry MS, Ryals B. Intranasal Ketamine for Abortive Migraine Therapy in Pediatric Patients: A Single-Center Review. Pediatr Neurol. 2020;104:46–53. doi:10.1016/j.pediatrneurol.2019.10.007

20. Petersen AS, Pedersen AS, Barloese MCJ, et al. Intranasal ketamine for acute cluster headache attacks-Results from a proof-of-concept open-label trial. Headache. 2022;62(1):26–35. doi:10.1111/head.14220

21. Sarvari HR, Baigrezaii H, Nazarianpirdosti M, Meysami A, Safari-Faramani R. Comparison of the efficacy of intranasal ketamine versus intravenous ketorolac on acute non-traumatic headaches: a randomized double-blind clinical trial. Head Face Med. 2022;18(1):1. doi:10.1186/s13005-021-00303-0

22. Harris PA, Taylor R, Minor BL, et al. The REDCap consortium: Building an international community of software platform partners. J Biomed Inform. 2019;95:103208. doi:10.1016/j.jbi.2019.103208

23. Hasan M, Hofstetter R, Fassauer GM, Link A, Siegmund W, Oswald S. Quantitative chiral and achiral determination of ketamine and its metabolites by LC-MS/MS in human serum, urine and fecal samples. J Pharm Biomed Anal. 2017;139:87–97. doi:10.1016/j.jpba.2017.02.035

24. Zanos P, Moaddel R, Morris PJ, et al. NMDAR inhibition-independent antidepressant actions of ketamine metabolites. Nature. 2016;533(7604):481–486. doi:10.1038/nature17998

25. Abbott JA, Popescu GK. Hydroxynorketamine Blocks N-Methyl-d-Aspartate Receptor Currents by Binding to Closed Receptors. Mol Pharmacol. 2020;98(3):203–210. doi:10.1124/mol.120.119784

26. Li L, Vlisides PE. Ketamine: 50 Years of Modulating the Mind. Front Hum Neurosci. 2016;10:612. doi:10.3389/fnhum.2016.00612

27. Moffa A, Costantino A, Rinaldi V, et al. Nasal Delivery Devices: A Comparative Study on Cadaver Model. Biomed Res Int. 2019;2019:4602651. doi:10.1155/2019/4602651

28. Suman JD, Laube BL, Dalby R. Comparison of nasal deposition and clearance of aerosol generated by nebulizer and an aqueous spray pump. Pharm Res. 1999;16(10):1648–1652. doi:10.1023/a:1011933410898

29. Suman JD. Current understanding of nasal morphology and physiology as a drug delivery target. Drug Deliv Transl Res. 2013;3(1):4–15. doi:10.1007/s13346-012-0121-z

30. Tashima T. Shortcut Approaches to Substance Delivery into the Brain Based on Intranasal Administration Using Nanodelivery Strategies for Insulin. Molecules. 2020;25(21). doi:10.3390/molecules25215188

